# Unequal starting lines: gendered barriers to participation and outcomes in a peer-led physical activity program for adolescents living with HIV in India

**DOI:** 10.64898/2026.08.24.26361285

**Authors:** Siddha Sannigrahi, Kacie Filian, Babu Seenappa, Hrishikesh Sathyamoorthy, Suhas Reddy, Meghana Gowda, Jeevitha Pushparaj, Ramu Sanju, Sahana Papanna, Satish Kumar SK, Michael Babu Raj, Lakshmi Ganapathi, Anita Shet

## Abstract

Adolescents with perinatally acquired HIV carry a high burden of depression and anxiety, and where specialist mental health services are scarce, peer-led physical activity has been proposed as a low-cost supportive intervention. Whether such programs reach girls and boys equally, and whether gendered constraints shape who is able to take part, has received little attention. Treating HIV status, gender, and adolescence as intersecting rather than additive axes of disadvantage, we examined participation in the Positive Running Program, a peer-led structured physical activity intervention delivered around antiretroviral therapy centers in Karnataka and Tamil Nadu, southern India.

We conducted a cross-sectional convergent mixed-methods study among 150 adolescents and young people with perinatally acquired HIV (100 boys and young men, 50 girls and young women; median age 17 years, interquartile range 15-19; 91% virally suppressed). Depressive and anxiety symptoms were screened using the Patient Health Questionnaire-9 and the Generalized Anxiety Disorder-7 scale, a score of 5 or above on either instrument was classified as a common mental disorder. High program adherence was defined as attendance at 65% or more of scheduled sessions. Associations were estimated using logistic regression adjusted for age, with gender-stratified models and an adherence-by-gender interaction term. Four focus group discussions with 28 participants and peer facilitators were analyzed using reflexive thematic analysis, with themes generated inductively and interpreted through an intersectional lens and through self-determination theory. Quantitative and qualitative findings were integrated at the interpretive stage.

Girls and young women attended fewer sessions than boys and young men (mean 61.6% versus 65.6%; p=0.025) and were less likely to reach the pre-specified ≥65% adherence threshold (10/50, 20% versus 57/100, 57%; p<0.001). They also had a higher prevalence of a positive depression screen (33/50, 66% versus 43/100, 43%; p=0.009) and of any common mental disorder (36/50, 72% versus 52/100, 52%; p=0.022). Higher adherence was associated with lower odds of a common mental disorder overall (adjusted odds ratio 0.31, 95% CI 0.13-0.68) and among boys and young men (0.33, 0.14-0.75); among girls and young women, only 10 participants met the adherence threshold and the estimates were imprecise. Qualitative findings located the constraints upstream of the program, in household authority over girls’ time, restrictions on mobility outside the home, care-giving obligations, and community disapproval of girls exercising in public.

The central finding concerns participation rather than benefit: girls and young women were half as numerous among participants and attended less consistently, clustering just below the high-adherence threshold. This differential opportunity to participate arises where gendered household authority intersects with the constrained autonomy of adolescence and the concealment demanded by HIV status. Interpreted through self-determination theory, the program supported competence and relatedness for those who attended but did little to secure the autonomy girls needed to attend consistently. The cross-sectional design precludes causal inference, including about the direction of the association between attendance and symptoms. Peer-led physical activity programs in this setting should treat gender inequality not as background context but as a determinant of participation and a core target of design.

## INTRODUCTION

India has the third-largest HIV epidemic globally with an estimated 2.3 million people living with HIV, and national antiretroviral therapy programs have achieved remarkable progress, resulting in an estimated 80% decline in HIV-related mortality since 2010.(1) India’s youth population is one of the largest globally; despite advancements in HIV care and support, adolescents account for 35% of new HIV infections annually.(2,3) Among groups that remain vulnerable are adolescents and young adults younger than 25 years of age with perinatally acquired HIV (APHIV), who navigate a period of rapid physical, emotional, and social change while managing a chronic, stigmatized illness.(4) Many reach sexual maturity without comprehensive HIV knowledge, increasing risks of onward transmission if they are not virologically suppressed.(5) The mental health burden among APHIV is substantial, with studies reporting depression rates up to 26% and generalized anxiety disorder in as many as 46%(6–9). These common mental disorders undermine adherence to antiretroviral therapy (ART), reduce viral suppression rates, and are linked to poorer health and social outcomes.(10–13) Reported prevalence varies substantially with the screening threshold applied: studies using a Patient Health Questionnaire-9 (PHQ-9) cut-point of 10 or above, indicating moderate or greater symptomatology, report considerably lower prevalence than those using a cut-point of 5 or above, which captures mild symptoms and is more sensitive to subclinical distress relevant to intervention targeting. For APHIV girls and young women, these challenges are further shaped by gender inequities rooted in deeply entrenched patriarchal norms. Such constraints are reflected in lower school retention, early marriage, and reduced opportunities to participate in supportive social and health-promoting programs.(14–17) Young women living with HIV may also face greater stigma, less access to healthcare, and higher levels of anxiety and depression than their male peers.(14–17) APHIV in India are often raised in residential childcare institutions, settings that structure daily routine, supervision, and permission to leave the premises in ways that differ substantially from family-based care. How gendered norms of supervision and mobility operate within these institutions, and how they shape girls’ access to health-promoting programs, remains largely undescribed. While prior research has described how gendered determinants intersect to shape HIV-related vulnerability among adolescent girls and young women in India, a critical gap remains in understanding how these determinants shape girls’ engagement with interventions, and how they operate differently across institutional and family-based living arrangements.(18)

Physical activity offers a promising, underutilized lifestyle-based pathway to improve both physical and psychological well-being in APHIV. Evidence from sub-Saharan Africa shows that sport-based interventions in youth can address mental health and social cohesion outcomes broadly, and that physical activity improves psychological status among adults living with HIV.(19,20) However, peer-led physical activity interventions are rarely designed with a gender-specific lens, leaving adolescent girls to often face unique sociocultural constraints, including restrictive gender norms, safety concerns, and lower encouragement for sports participation compared to boys. (21) In India’s context, culturally adapted gender-responsive physical activity interventions, particularly for APHIV, remain scarce.

To support health and social outcomes among APHIV, a peer-led physical activity intervention called the Positive Running program was initiated in southern India in 2022, reaching over 200 young APHIV and those affected by HIV in the southern Indian states of Karnataka and Tamil Nadu. This study examined gendered patterns of participation in the program and the relationship between participation and mental well-being. We asked, first, whether adolescent girls and boys participated at comparable levels; second, how participants and peer implementers themselves accounted for any difference; and third, whether more consistent participation was associated with a lower burden of depressive and anxiety symptoms. The evaluation was conducted using a community-based participatory design in which trained youth investigators living with HIV co-led instrument development, data collection, and analysis. We used intersectionality as an orienting framework for understanding how gender, HIV status, and adolescence operate together rather than additively, and self-determination theory as an interpretive lens for participants’ accounts of what sustained or undermined their engagement. The frameworks were not operationalized through validated measurement instruments, instead they were both applied to interpret qualitative findings generated inductively. We found a substantial and consistent gender gap in participation, together with qualitative accounts that locate its origins in household and community constraints operating before adolescents reach the program.

## METHODS

### The intervention: Positive Running

The *Positive Running* program is a peer-led physical and psychosocial intervention designed for adolescents and young people living with or affected by HIV. The program integrates structured physical activity with resilience-building and leadership development to promote overall health and well-being. The intervention has a weekly schedule consisting of two strength and conditioning sessions and two running sessions. Strength and conditioning includes resistance, cardiovascular, and flexibility exercises to improve muscular strength, endurance, and mobility, respectively. Each running session targets age-appropriate individual performance goals: threshold runs emphasize sustained moderate effort to build endurance; speed runs focus on short, high-intensity intervals to enhance power; and distance runs aim to increase aerobic capacity and mental resilience. In addition to physical training, participants attend quarterly educational camps covering topics such as nutrition, exercise physiology, hygiene, and injury prevention. These camps also emphasize resilience-building, teamwork, and goal setting to promote holistic development and psychosocial well-being. Participants also take part in at least one regional or national running event, providing opportunities to apply their training and build confidence. These activities are led by youth peer implementers called captains, who undergo training to serve as intervention implementers. They lead exercise sessions, guide participants through the program, and foster a supportive, motivating environment. Working closely with senior program leadership, Captains help maintain program consistency, encourage participation, and ensure the intervention’s overall success and sustainability.

### Community-based participatory research

Impact evaluation of the intervention was conducted through a community-based participatory research (CBPR) approach, with youth living with HIV actively engaged throughout the research process (youth investigators).(22) Five trained youth investigators were part of the research team. Their contributions were essential in refining surveys and interview guides to improve comprehension, strengthening the study’s cultural and contextual validity, and ensuring that research activities reflected the lived realities of youth with HIV in Karnataka. All youth investigators completed Human Subjects Research certification through Johns Hopkins University and underwent a 2-day training in sensitive and ethical survey administration and qualitative research techniques.

### Study setting and participants

Between March and April 2024, we enrolled participants from childcare institutions located in Bijapur, Dakshina Kannada, and Kolar districts in Karnataka, and in Krishnagiri district in Tamil Nadu. These institutions provide residential care, psychosocial support, and educational or vocational training for children and adolescents <18 years of age living with or affected by HIV. Children in non-institutional settings and young adult participants living with their respective families were enrolled from Bangalore and Bijapur. Study eligibility criteria included being (i) between 12 to <25 years of age; (ii) currently enrolled in the *Positive Running* program for at least 12 months; (iii) having documented HIV status prior to 10 years of age, or HIV-affected status (defined as having one or both parents with HIV-confirmed status).

### Ethical approval and consent

The study was approved by Institutional Review Boards of the Y.R. Gaitonde Centre for AIDS Research and Education (YRGCARE), Chennai, India (#YRG375), and the Johns Hopkins Bloomberg School of Public Health, Baltimore, USA (#IRB00023077). Written informed consent was obtained from participants ≥18 years. For participants <18 years, written informed consent was obtained from a parent (if available), legal guardian, or the legally authorized representative of the childcare institution, and oral assent was obtained from the participant in the presence of a child advocate. Assent discussions were conducted privately, away from institutional staff and peers, and participants were told that declining to participate would not affect their care, their standing in the institution, or their continued enrolment in the Positive Running program. Conversations took place in the participant’s preferred language to ensure comprehension and voluntary participation. All procedures conformed to the Declaration of Helsinki.

### Study design and indicators

We employed a convergent parallel mixed-methods design incorporating quantitative and qualitative research. Following informed consent, trained youth investigators administered surveys in person, with measures in place to ensure privacy and confidentiality during data collection. These measures included conducting surveys in a private setting away from peers and staff, ensuring that responses could not be seen by others, and securely storing completed forms in locked folders accessible only to authorized research staff. To limit social desirability and reporting bias, surveys were administered privately by trained youth investigators rather than by program implementers, participants were assured that responses would not affect their standing in the program, and clinical indicators were verified against medical records where available.

i. Survey: Survey components included sociodemographic details such as age, gender, education, and residential history. Gender was self-reported by participants as male or female; no participant was identified under another category. We use the term gender throughout rather than sex, as our analytic interest is in socially constructed roles. Clinical information such as key HIV-related indicators from the past 12 months, including hemoglobin, CD4 cell count, viral load, and current antiretroviral therapy (ART) regimen was obtained through participant self-report and medical records. Psychosocial health status was assessed using the 9-item, Patient Health Questionnaire (PHQ-9) and 7-item, Generalized Anxiety Disorder (GAD-7) to screen for depressive and anxiety symptoms.(23,24) Each tool uses a 0-3 symptom-frequency scale, with PHQ-9 scores ranging from 0-27 and GAD-7 scores from 0-21. A positive screen for any depression was defined as PHQ-9 scores ≥5 (mild, moderate, or severe), while scores 0-4 was categorized as no depression. A positive screen for any anxiety was defined as GAD-7 scores ≥5 (mild, moderate, or severe), with scores 0-4 categorized as no anxiety. A pre-specified safety protocol governed responses to the mental health screening tools; any significant response (i.e., item 9 ≥1, PHQ-9 ≥15, or GAD-7≥15) triggered immediate notification of the onsite counsellor, who conducted a same-day risk assessment in private and, where indicated, initiated referral to the affiliated tertiary care center. Youth investigators were trained in this escalation procedure.
ii. Intervention adherence: The *Positive Running* intervention adherence was monitored in real-time by Captains. Daily attendance was documented on site and was quantified as a percentage of days attended among all required days of the program minus holidays, calculated over the full duration of each participant’s enrollment in the program. Based on program benchmarks, a pre-specified attendance of ≥65% was considered as ‘high adherence’ to the intervention.
iii. Qualitative research: We conducted focus group discussions (FGDs) and in-depth interviews (IDIs) in-person using purposive sampling from amongst the survey participants. Of 35 individuals invited to take part in FGDs, seven declined, citing reasons of school or employment commitments. These qualitative methods aimed to identify motivations for participation in the PRP, perceived benefits, and barriers to sustained engagement, with particular attention to gender-related factors influencing participation and psychosocial outcomes. Four FGDs (n = 28; boys and young men=14, girls and young women=14) were conducted across four participating childcare institutions. Each discussion group consisted of 6-10 participants of the same gender and age range (13-15 years or 16-19 years). FGDs were co-facilitated by gender-concordant youth investigators trained in focus group moderation, who ensured cultural and linguistic appropriateness. We also conducted four IDIs (n = 4; young men=2, young women=2) with current program implementers (Captains) who were also former *Positive Running* participants to further contextualize and deepen understanding of participant experiences. All four Captains approached for in-depth interviews agreed to participate. Sessions lasting approximately 30-60 minutes were conducted in English and Kannada in a private setting to ensure privacy and confidentiality. With participants’ consent, discussions were audio-recorded. No repeat interviews were conducted. Data sufficiency was assessed using the concept of information power rather than saturation alone. Researchers conducting the FGDs and IDIs were SS, KF (both female) and MBR (male), with Masters and PhD in public health research, and experienced in conducting qualitative research. Co-facilitaters included youth investigators (BS, SR, MG, JP), who were part-time mentors and students at the time of the study and held no formal research qualifications. No institutional staff, program implementers, or other non-participants were present during the discussions. Facilitators recorded brief field notes during and immediately after each session, documenting group dynamics, non-verbal responses, and contextual observations; these informed interpretation during analysis but were not formally coded. Transcription and translation from Kannada were conducted by bilingual members of the research team; and a subset of transcripts was back-checked against audio recordings by a second bilingual team member. Formal member checking was not undertaken, although youth investigators contributed to interpretation of findings.

## Data Analysis

### Quantitative analysis

Participant characteristics, clinical indicators, intervention adherence, and mental health screening results were summarized descriptively and disaggregated by gender (Table 1). Continuous variables were reported as medians with interquartile ranges and categorical variables as counts with percentages, with numerators and denominators shown. Analyses were conducted in R version 4.3.2, using two-sided tests with alpha set at 0.05. Two-sided Fisher’s exact tests assessed associations between gender and categorical outcomes, including positive screens for depression and anxiety, viral suppression, and high intervention adherence.(25) Logistic regression models estimated unadjusted and age-adjusted prevalence odds ratios for a positive screen for common mental disorder in relation to high adherence. To assess effect modification by gender, we fitted a multiplicative adherence-by-gender interaction term in the age-adjusted model; gender-stratified age-adjusted models are additionally presented for descriptive purposes, and are not interpreted as evidence of differential effect in the absence of a significant interaction. To assess the influence of the high adherence threshold, we repeated the age-adjusted model using alternative thresholds of 60%, 75%, and 80% attendance. Among girls, only 10 met the high-adherence threshold, yielding a stratified estimate with a confidence interval too wide to support inference in either direction; this estimate is reported for completeness and is not interpreted. As all eligible enrolled participants were included, no a priori sample size calculation was performed; the study was not powered for gender-stratified estimation, and stratified estimates are therefore exploratory. Analyses were complete-case, and the number contributing to each model is reported in Table 2.

**Table 1.**
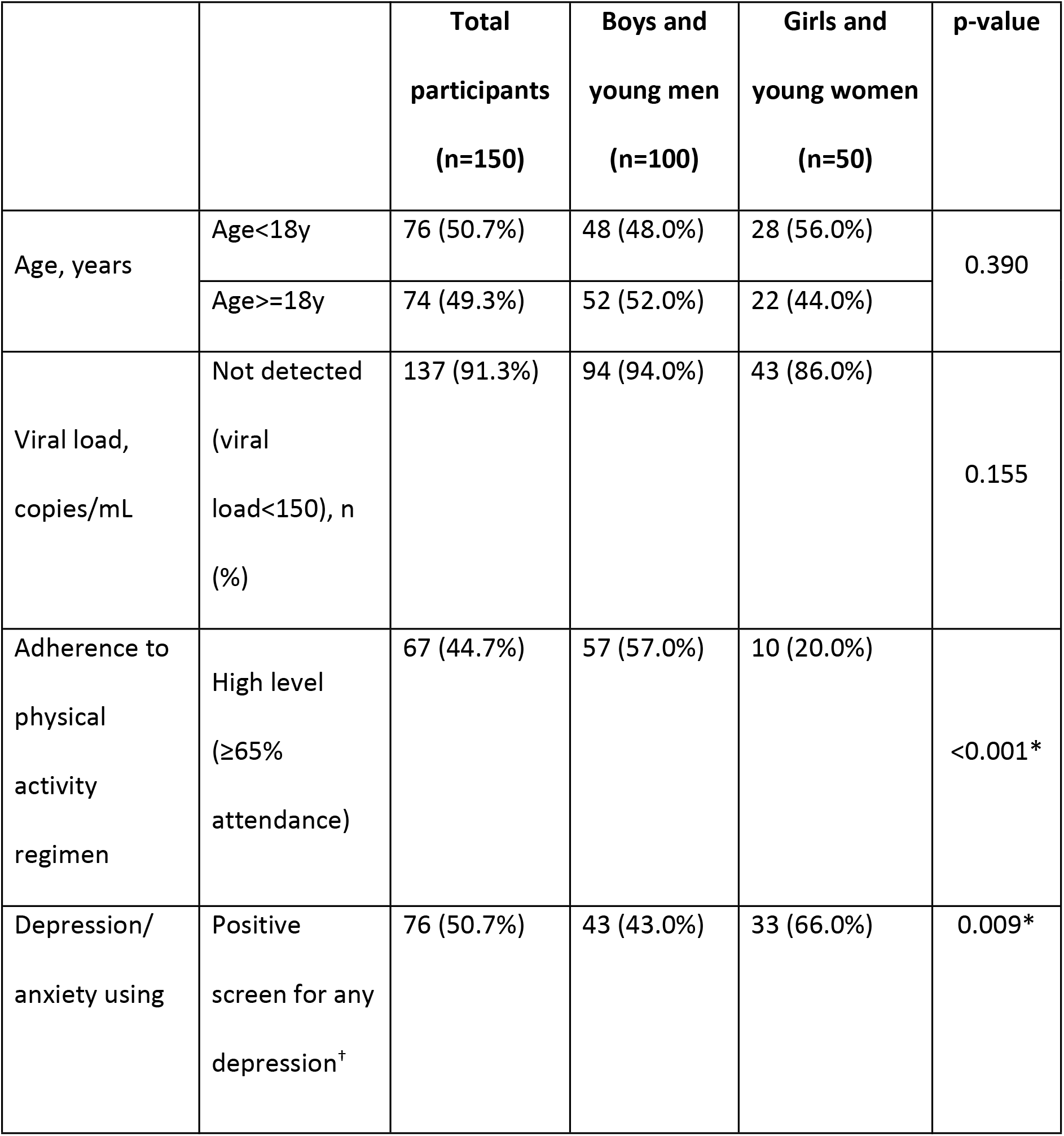
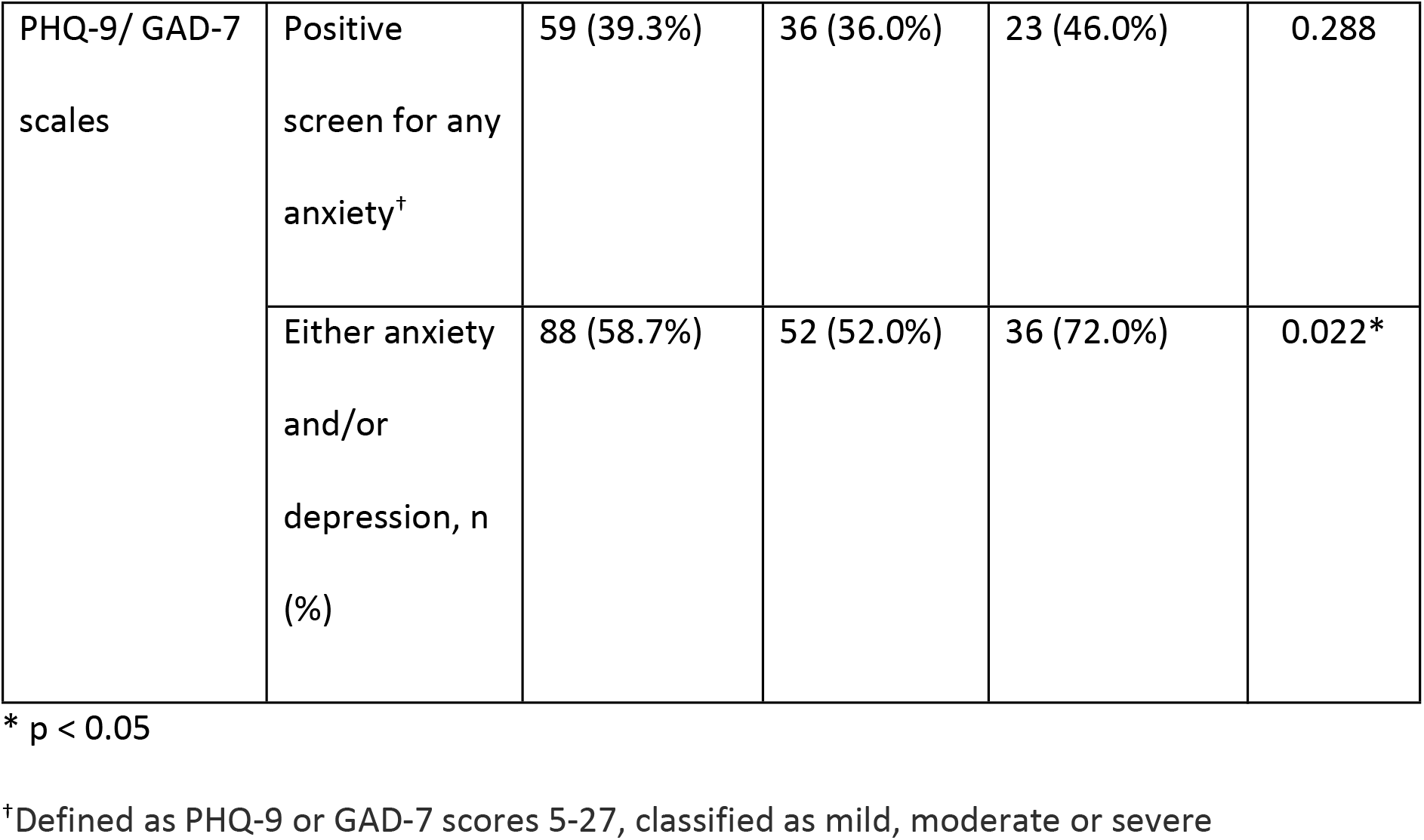
Gender-disaggregated sociodemographic characteristics (gender, age, viral suppression), intervention adherence, prevalence of depression and generalised anxiety disorder (GAD) among participants.

**Table 2.** Association between intervention adherence and common mental disorder (reference category: low intervention adherence (<65% attendance)

| Analysis | Odds ratio for prevalence of common mental health disorder<sup>a</sup> (95% CI) |
| --- | --- |
| **Overall** |  |
| Unadjusted | 0.44 (0.23-0.85)* |
| Adjusted for age | 0.31 (0.13-0.68)* |
| **Gender-stratified analyses<sup>†</sup>** |  |
| Girls and young women (n=50) | 4.33 (0.50-37.92) |
| Boys and young men (n=100) | 0.33 (0.14-0.75)* |

\* p < 0.05
† Gender-stratified models are adjusted for age.
<sup>a</sup> Common mental disorder is defined as a positive screen (mild severity or above) for anxiety and/or depression using the GAD-7 and PHQ-9 scales.

### Qualitative analysis

We analyzed data using reflexive thematic analysis, guided by Braun and Clarke’s six-phase framework.(26) Two researchers independently generated initial codes across English transcripts using Dedoose v9, using an inductive approach to identify patterns of meaning within participant narratives. Independent coding was used to broaden interpretive range rather than to establish reliability; no inter-coder agreement statistic was calculated, as reflexive thematic analysis does not treat coding convergence as a quality criterion. Codes were discussed collaboratively and refined, with attention to how gender shaped participants’ experiences of physical activity. Related codes were then organized into themes, and then further defined and named through team discussion and interpreted in relation to self-determination theory and intersectionality. Participant attributions denote self-reported gender, and are abbreviated for brevity in tables and quotes.

### Reflexivity

The research team comprised investigators based in India and the United States, and five youth investigators living with HIV. Coding was led by a US-based and an India-based researcher, both with epidemiological training and neither sharing participants’ lived experience of HIV. Youth investigators and India-based co-investigators led interpretation of culturally situated accounts, including those concerning family authority and community expectations. Youth investigators who co-facilitated the focus groups were known to some participants through prior involvement in the Positive Running program, and Captains interviewed as key informants were themselves former participants. Facilitators introduced themselves at the outset, explained their role in the research and their own connection to the program, and emphasized that participants’ responses would not be shared with program staff or affect their standing in the program.

### Theoretical frameworks

Themes were generated inductively and subsequently interpreted through two complementary frameworks(27,28). Intersectionality informed our reading of how gender and HIV status operate together rather than additively, compounding stigma and producing layered constraints on participation.(27) Self-determination theory, which holds that sustained motivation for health behaviour depends on the satisfaction of three psychological needs: autonomy, or perceived control over one’s decisions; competence, or self-efficacy in achieving valued outcomes; and relatedness, or social connection and belonging, provided a vocabulary for participants’ accounts of what supported or undermined their engagement.(28) Neither framework was operationalized through validated measurement instruments; both were applied interpretively to situate participants’ narratives within established theory.

### Integration

Quantitative and qualitative strands were analyzed independently and integrated at the interpretive stage, with qualitative findings used to explain the patterns of participation observed in the quantitative data.

## RESULTS

### Survey results

Of the 172 participants who completed 12 months of the *Positive Running* intervention, 155 had HIV status confirmed prior to 10 years of age (defined as APHIV), and were eligible for this analysis. Participants who were HIV-affected but not living with HIV (n=17) were excluded from this analysis. Among eligible participants, 150 (96.8%) completed the survey; five did not, owing to transfer (n=2), program discontinuation (n=2), and illness at the time of assessment (n=1). Median age was 17 years [Interquartile range (IQR): 15-19 years] and 100/150 (66.7%) were boys and young men and 50/150 (33.3%) were girls and young women. Viral suppression (VL <150 copies/ml) was documented in 137/150 (91.3%) of participants with no significant difference by gender (94.0% vs 86.0%, p=0.155). Mean intervention adherence across the program period was 64.3% overall (SD 10.7) (Table 1). Girls and young women attended fewer sessions than boys and young men (mean 61.6% versus 65.6%; p<0.05) and were substantially less likely to reach the pre-specified ≥65% adherence threshold (10/50, 20% versus 57/100, 57%; p<0.001), reflecting a female distribution concentrated just below that cut-point of 65% adherence (Figure 1).

**Figure 1.**
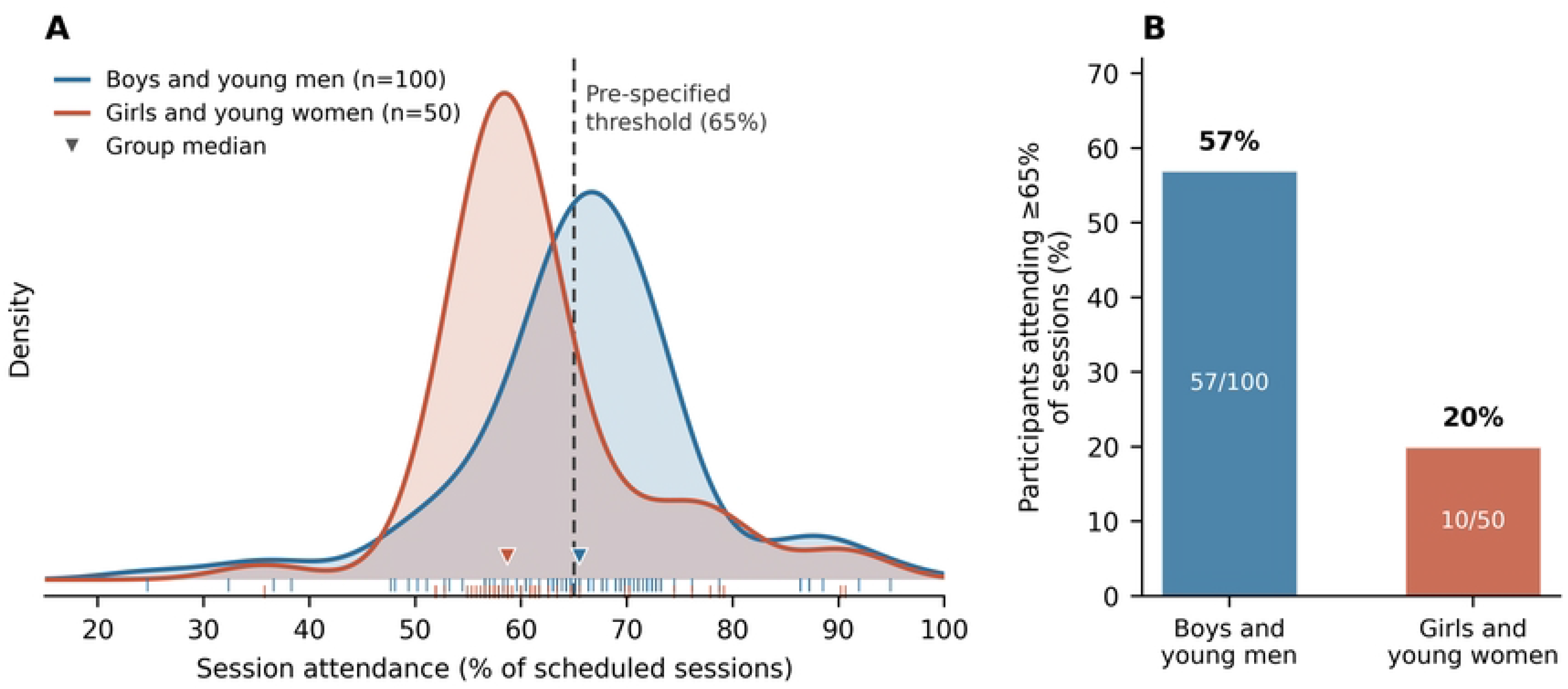
Program adherence by gender

Over half of participants (58.7%) screened positive for at least one common mental health disorder (depression and/or anxiety); 50.7% screened positive for depressive symptoms on the PHQ-9, and 39.3% screened positive for anxiety on the GAD-7 scale (defined as mild and above category based on standard scoring procedures for PHQ-9 and GAD-7). Gender-disaggregated estimates revealed a significantly higher prevalence of a positive screen for depression and at least one common mental disorder among girls and young women compared with boys and young men; 66% girls versus 43% boys met screening criteria for depression alone (p = 0.009), and 72% girls versus 52% of boys met screening criteria for a positive screen on either instrument (p=0.022). (Table 1). Anxiety screening did not differ significantly by gender (23/50, 46.0% vs 36/100, 36.0%; p=0.288).

High adherence to the intervention was associated with a lower odds of a positive screen for a common mental health disorder (unadjusted OR 0.44, 95% CI 0.23-0.85), with a larger estimate after adjustment for age (aOR = 0.31, 95% CI: 0.13-0.68) (Table 2). The adherence-by-gender interaction term was not statistically significant, providing no formal evidence that the association differed between young men and women. In gender-stratified age-adjusted models, the estimate among boys was consistent with the overall association (aOR 0.33, 95% CI 0.14–0.75). Among girls and young women, only 10 of 50 participants met the high-adherence threshold; the resulting estimate was imprecise, with very wide confidence intervals (aOR 4.33, 95% CI 0.50–37.92). We report this estimate for completeness and do not interpret it, as the girls’ stratum contained too few high-adherent participants to support inference in either direction.

Sensitivity analysis using alternative adherence thresholds of 60% and 75% yielded directionally consistent overall estimates; at 80%, the association was no longer evident, reflecting sparse data in both strata.

### Qualitative results

Thematic analyses from FGDs and IDIs explored the gendered, cultural, and structural factors shaping participants’ engagement in physical activity. The first section describes barriers to female participation, interpreted through an intersectional lens attentive to how gender, age, and HIV status operate together to constrain engagement both within and outside institutional settings (Table 3). The second section describes facilitators of participation, interpreted through self-determination theory, and includes accounts of shifting gender attitudes among boys that emerged unprompted across groups (Table 4).

**Table 3.** Themes of gender-related barriers contextualized using intersectional theory and participant quotations from focus group discussions and in-depth interviews.

|  |  |
| --- | --- |
| Themes | FGD/IDI Quotations |
| **Gendered roles and responsibilities** | *They are never rewarded for their efforts, only boys are hired for any work, and girls have an attitude that they don't know anything. - Female, Participant*<br>*[They say], if you run outside like men, what will others think when seeing you? - Female, participant* |
| **Stigmatisation of women in sport** | *Many girls face discrimination because of the running and exercising...I remember people saying, 'you're learning very dirty things'. - Female, participant*<br>*That's the comment they get in the villages, 'if you run, what will others think?' - Female, Captain* |
| **Cultural expectations of modesty** | *Before when they would go and participate in their school runs, teachers would not take them for running because they thought they were weak and can't run. - Female, Captain*<br>*They all come from the villages background and all, and some families know that running will help their health. But some people are not educated about this, so they think wearing a sport shirt is very bad, so they don't allow it worn in their homes. - Female, Captain* |
| **Lack of decision-making power** | *No, they're not allowing me to do exercise, saying 'You're a girl, what do you want to do? Simply sit in home, do homework and stay here. No need to run outside. - Female, Captain*<br>*People think that nothing will happen in our hands. We must go beyond their words. - Female, participant* |

**Table 4.** Themes of motivation contextualized using Self Determination Theory and participant key words and quotations from focus group discussions and in-depth interviews.

| Theme | Subthemes | Key Words | Illustrative Quote |
| --- | --- | --- | --- |
| **Autonomy** | Self-managed health<br>Leading and learning sessions during camps<br>Gender norms challenged with action | *“fit”*<br>*“strong”*<br>*“control”*<br>*“responsibility”*<br>*“equal”* | *Many younger peers were so weak and having so much of cough and all.*<br>*So, after they started running and exercising, it has reduced. So many have shared with me, ‘Before regularly I was getting cough and all, but after joining the run and doing running exercise my cough has reduced. I’m*<br>*becoming strong. I feel this strength.'*<br>*They want to continue to be healthy, so they run.*<br>- Female, Captain |
| **Competency** | • Health knowledge<br>• Confidence built through distance milestones<br>• Recognition and achievement | *"improvement"*<br>*"achievement"*<br>*"capable"*<br>*"goals"*<br>*"better"* | *When they started, they were not doing much. Most of the peers were telling me that it's difficult to run saying, 'I cannot run'. Then, we slowly pushed them to do 2 kilometers, then 5 kilometers. Then they say, 'we feel that we can do more than before, like we can do better, more than other children... because some children don't get opportunity to do, but we get to.*<br>- Female, Captain |
| **Relatedness** | • Peer bonding through team runs | *"share"*<br>*"together"*<br>*"team"* | *Like if I run alone, I don't feel any motivation because I'm all by myself.*<br>*But when we come together for the* |
|  | • Collective motivation through Captains' mentorship<br>• Increased sense of belonging fostered through mentoring and being mentored | *"inspire"*<br>*"encourage"* | *camps, we get many friends. We have lots of fun. We all will jump in and we will have lots of fun. So that's the key motivation I think.*<br>- Male, Captain |

#### ***I.*** Barriers to girls’ participation in physical activity

Participants and peer implementers (captains) described four interconnected constraints on girls’ participation: gendered roles and responsibilities, stigmatization of women in sport, cultural expectations of modesty, and limited decision-making authority. (Table 3).

### Gendered roles and responsibilities

Participants described encountering negative responses from family and community members when they attempted to exercise, whether within or outside the institution.

> *‘So the parents feels that our girls have to be a certain way only and they should not do boys’ work. Girls can only do girl work and homework, so they should not do running and other things. They have to be inside the home and work.’* - Male, CaptainThese accounts reflect prevailing norms in which outdoor physical activity is regarded as inappropriate for girls and women, whose time is expected instead to be given to household responsibilities. Framing exercise as a male activity was described as discouraging girls’ participation directly, through explicit prohibition, and indirectly, by positioning running as something girls had no legitimate claim to.

### Stigmatization of women in sport

Participants described a pattern of social disapproval directed at girls who exercised in public. This took an active form in comments from neighbors and community members, with running characterized as unclean or improper. Participants described the resulting fear of judgment and social exclusion as a substantial disincentive to continued participation.

> *‘That’s the one thing; they talk bad about the girls. Because of that, they stop running or they don’t have any interest or confidence to do it’* - Female, Captain

### Cultural expectations of modesty/femininity

Constraints at the community level centered on expectations that young women present themselves modestly in public. Participants reported that clothing suited to running, such as shirts and athletic trousers, drew criticism from community members and was in some cases prohibited by family members when girls left the house to exercise. One participant described running in her village as constrained less by the physical demands of the activity than by anticipated scrutiny and the possibility that her family’s standing would be questioned.

> *‘We feel that we have done something wrong. No one is interested in doing it again if they say no. What will [they] talk again, what will [they] say?’*- Female, participant

### Limited decision-making authority

Participants described having little authority over decisions about leaving the institution or the household to exercise. Accounts located this authority with adult male household heads, with several participants describing personal motivation to continue running that could not be acted upon without permission. These accounts were articulated primarily in terms of gender and age: participants described constraint as something they experienced as girls and as young people, rather than as something they attributed to living with HIV. The intersection with HIV status was less explicitly named. It is plausible that the concealment and social caution that HIV status demands in these communities compounds the surveillance participants described, and prior work in India documents the layered stigma faced by women and girls living with HIV; however, our data do not allow us to establish this directly, and we present the intersectional reading as an interpretive frame rather than as a finding grounded in participants’ explicit accounts.

> *‘We see in many villages there are many families they don’t send their girls anywhere outside, especially to work or study in different places apart from their village. They won’t allow girls, or they won’t encourage them to run.’* - Female, Captain

#### ***II.*** Facilitators of participation

Participants’ accounts of what sustained their engagement mapped onto the three psychological needs described by self-determination theory: autonomy, competence, and relatedness (Table 4). Participants described gains they attributed to sustained participation, extending in some accounts beyond physical activity to confidence and educational engagement. These are participants’ own attributions rather than measured outcomes, and we report them as such. Accounts in this section come from both girls and boys, and from participants and peer implementers; attributions are noted for each.

#### Autonomy

Participants described exercising greater choice over health-related behaviours and attributed changes in their health to their own decisions rather than to instruction from staff. Accounts included fewer episodes of illness, greater emotional steadiness, and enjoyment of the activity itself.

> Last year there was no exercise, so I fell sick two or three times, this time I did not fall sick even once because of running and exercise. - Male, participant

Female peer implementers additionally described increases in independence that extended beyond the program, including travelling greater distances alone, managing finances, and taking on responsibility for training others.

> *I feel more confident in me because I have learned so many things. Now it may be travelling, it may be spending money. It may be training others, talking to others, like giving them suggestions. It has improved many things in my life, and I can also take up many responsibilities. It has mainly increased my confidence and especially increased my confidence in myself. Now I can trust myself so much. -* Female, Captain

Some girls described responding to community disapproval by continuing to participate anyway, framing their health as a matter within their own remit rather than one subject to community judgment.

> *Let them think what they want, we have to take care of our own health matters.* - Female, participant

### Competence

Participants described growing confidence in their physical capabilities and in their health knowledge. Nutritional and hygiene sessions were described as giving participants information they could act on and share with peers.

> *We know that when we are running and exercising, we need to have good nutritious food daily. Like if we are getting groggy, to eat more egg or any fruits and vegetables. So, we have to have our daily nutritious food on time and mainly take your tablets on time. -* Female, Captain

Participants also described progressive distance goals and participation in organized runs as experiences of mastery, and connected these to confidence in other areas, including school engagement and plans for future employment.

> *There are any running competitions in our school, if we run well here and practice well, we can go there and run well. If we run elsewhere, our school will get a good name and our teacher will also be happy. -* Female, participant

#### Relatedness

Participants described a sense of belonging and mutual support arising from shared activity, and identified running camps in particular as occasions on which sustained friendships formed.

> *Running with new people and participating and talking with new people, it’s very nice. We can share how we are feeling while running. What are our difficulties. We can share all those things. All are challenges about running while we are running and all. So, it will be good to share each other’s opinion. So, we can get to know each other better and we can keep in contact with them. -* Female, participant

Peer implementers’ presence during institutional visits was described as offering an opportunity to raise concerns and as a source of encouragement to continue participating when girls encountered gender-related obstacles.

> *They see me, and they’ll also start doing and they’ll be also saying, ‘I want to be like you, I want to do this exercise like you’ and they say, ‘How are you doing these many things?’. So I say that I learned from the others and I’m teaching for them. Then I say, ‘you have to learn from me, and you have to teach for others’. -* Male, Captain *Courage comes when [Captain] is with us. -* Female, participant

#### Shifting attitudes among boys toward female participation

A theme that emerged across groups without prompting concerned changes in how the males regarded females’ physical capabilities. Young men who trained alongside young women at mixed camps described observing females completing the same distances, and connected this to a revised view of what females were capable of. Participants and peer implementers alike framed shared training as the mechanism, describing camps as occasions on which capability became visible across a divide that institutional separation ordinarily maintained. Boys and young men described how seeing girls and young women excelling in sports challenged preconceived, socially constructed differences in abilities across genders (i.e. girls can’t run as far or as fast, boys lead physical activities while girls only socialize).

> *Nowadays boys and girls are at different institutions. So, when they come back together for a camp, they can see each other’s capabilities. Like sometimes boys may be thinking that, ‘Yeah, we are good at something’ and they may not know that there’s a girl better than them at it. So, when they come together, they can understand other’s abilities so that they can be inspired and they can change their mindset that girls can do it too. -* Female, Captain

> *I want them [girls] to run forward like us. -* Male, participant

Girls and young women, in turn, described visible female peer implementers as roles models for what they might themselves achieve. We note that these accounts describe stated attitudes at a single point in time; we did not measure attitudes before and after participation, and cannot establish whether or for how long such shifts persist.

> The *program is showing even the girls can run 5K and 10K. It is an example for the girls that you can also play any games, and you are equal to the boys. -* Male, Captain

#### Divergent cases and minor themes

Not all accounts conformed to the dominant pattern of constraint. A minority of girls and young women described continuing to participate in the face of community disapproval, framing health as a matter within their own control rather than one subject to community judgment, and rejecting the authority of those who objected. These accounts were fewer in number than accounts of accommodation to gendered expectations, and were concentrated among older participants and peer implementers rather than younger participants. A second minor theme, shifting attitudes among boys and young men toward female participation, arose unprompted across all groups and ran counter to the expectation that mixed-gender activity would reinforce existing norms. Both were retained in the analysis rather than treated as outliers, as they identify conditions under which the constraints described were partially loosened.

## DISCUSSION

This mixed-methods evaluation of a peer-led physical activity program for adolescents and young people with perinatally acquired HIV in southern India produced three principal findings. First, girls and young women participated far less consistently than boys and young men: only a fifth reached the high-adherence threshold, compared with well over half of boys, and girls carried a substantially higher burden of depressive and anxiety symptoms. Second, participants located the reasons for this gap upstream of the program itself - in household authority over girls’ time and movement, community surveillance of girls’ conduct and appearance, and expectations that girls’ hours belong to domestic work. What girls and young women described lacking was not motivation but the authority to act on it; several spoke of wanting to continue running and being unable to decide for themselves whether they could. Third, and least anticipated, the program appeared to shift how boys understood girls’ capabilities. Young men and boys who trained alongside young women and girls at mixed camps described watching females complete comparable distances and revising assumptions they had held about what females could do. This was an account echoed by girls, who described visible female peer implementers as evidence of what was possible for them. Taken together, these findings shift the question the program poses. The equity problem here is not that girls participated and benefited less; it is that most girls could not participate consistently in the first place. Yet the same program, for those it did reach, generated exactly the kind of shared, visible achievement that appears capable of loosening the norms constraining participation. That the intervention both ran up against gendered constraint and, in a small way, began to work on it is the central tension this paper describes.

### Health and psychosocial gains

Physical activity has been associated with improved psychological wellbeing among people living with HIV through physiological, psychological, and social pathways.(29,30) Much of this evidence derives from adult populations and from designs able to establish temporal ordering; our cross-sectional data cannot do so. Adolescents in this cohort who attended more consistently described reduced fatigue, greater emotional steadiness, and increased self-esteem, and had a lower prevalence of positive mental health screens. Two interpretations are consistent with these data: that participation supported psychological wellbeing, or that adolescents experiencing less psychological distress were better able to attend consistently.(31,32) Longitudinal evidence from general adolescent populations suggests the relationship is bidirectional, and both processes may operate here.(33) What our data establish more securely is that low-cost, community-embedded physical activity programming is feasible in this population and setting, and that adolescents themselves describe it as valuable.(34,35)

### Peer-led model and self-determination

The program’s peer-led structure emerged in participants’ accounts as central to sustained engagement. Captains, trained peers with lived experience of both HIV and the program, were described as relatable models who normalized physical activity and made it appear achievable. Participants’ accounts of what sustained their engagement corresponded to the three needs described by self-determination theory: control over health decisions, growing confidence in physical and health-related competence, and a sense of belonging generated through shared training(28,36). While these mechanisms promote immediate improvements in emotional regulation and well-being, they also underpin sustained adherence to care and long-term psychosocial resilience. We did not measure these constructs with validated instruments, and offer this correspondence as an interpretive reading rather than a test of the theory. The reading is nonetheless consistent with evidence that need-supportive environments are associated with sustained physical activity across settings, and suggests that peer leadership may be an active ingredient rather than an implementation convenience.(36) Whether such engagement persists beyond the program period cannot be determined from these data.

### Gendered pathways to physical activity adherence and well-being

In India, deep rooted expectations around modesty, safety, and domestic responsibilities restrict women’s mobility and ability to participate in physical activity.(37–39)This is embedded in the perception of physical activity as a symbol of undesirable modernity, a rejection of traditional values that can invite social disapproval and harassment, to which young women are particularly vulnerable.(40–42) Even when women succeed in navigating these sociocultural barriers, they continue to face gender bias in the allocation of funding, sports infrastructure, and training opportunities, which disproportionately favor men.(41) These structural inequities are reflected in gendered differences in mental health in this cohort. Prior research across diverse cultural contexts, identifies constraints on women’s participation in sport operating across physiological, psychological, sociocultural, and environmental.(41,42) All four were represented in participants’ accounts here, with sociocultural constraints (community disapproval, expectations of modesty, and limited decision-making authority) the most prominent, indicating that even within institutional settings, gendered expectations shape engagement.

These findings extend existing literature by demonstrating how these constraints operate in a population also managing a stigmatized chronic infection.(37–42) Prior work describes how adolescent girls’ mobility in India is curtailed by social surveillance and moral policing, processes that intersect with HIV-related stigma.(15,18) Notably, participants in our study articulated constraint predominantly in terms of gender and age rather than HIV status. This may reflect the institutional setting, where HIV status is shared among residents and therefore less salient as a marker of difference, or it may reflect the difficulty of naming a stigma one has been taught to conceal. Either reading is consistent with an intersectional account, but our data do not adjudicate between them.

The marked disparity in program adherence where over half of young men achieved high attendance compared to only one-fifth of young women, is the clearest finding of this evaluation, and the qualitative data offer a coherent account of it. Girls and young women’s psychological needs were often undermined by external restrictive gender norms and cultural expectations of modesty. Girls identified a lack of decision-making power and social surveillance as primary barriers, where the patriarchal head of the household often dictated their mobility and ability to exercise outside the home. These are constraints operating before an adolescent reaches the program, and no feature of program design encountered at the point of delivery can fully offset them. We are unable to determine whether physical activity confers comparable mental health benefit for girls as for boys in this population. Only 10 girls attended consistently, and the resulting stratified estimate is uninformative, and it would be misleading to conclude that participation does not benefit girls.

Several mechanisms could plausibly produce genuinely attenuated benefit among girls, were a sufficiently powered study to find it. Girls and young women with HIV face a heavier burden of concurrent stressors, including stigma, caregiving responsibilities, restricted autonomy, and exposure to violence, any of which might offset gains from physical activity.(14–16) Alternatively, drawing on self-determination theory, participation that is externally negotiated or permitted rather than self-directed might support competence and relatedness while leaving autonomy unaddressed, and thus yield smaller psychological returns. We raise these as hypotheses for adequately powered longitudinal work, not as interpretations of the present estimates.

### Gender-transformative approach of the Positive Running model

Several program features appear to address gender-related constraints at the point of delivery. Supervised, progressive training reduces concerns about injury and physical capability; a non-competitive, peer-led structure creates an environment in which athletic clothing and visible exertion attract less judgment; and organized group activity offers a socially sanctioned context for girls’ exercise in communities where independent female mobility is constrained. These features may explain why some young women sustained participation despite considerable opposition. They did not, however, produce equitable participation. That the gap persisted at this magnitude within a program designed to be inclusive suggests that constraints operating at the household and community level are not substantially modifiable through program design alone. Systematic reviews of gender-transformative programming reach a consistent conclusion: interventions that engage the gatekeepers of girls’ participation such as parents, male household heads, community leaders, achieve more than those that work only with girls themselves.(43,44) One finding does point toward transformative potential. Boys who trained alongside girls at mixed camps described revising their assumptions about girls’ physical capabilities, framing their completion of comparable distances as evidence of equal capacity. Engaging boys and young men as participants in norm change, rather than as obstacles to it, is increasingly recognized as a component of effective gender-transformative programming. Our data capture stated attitudes at a single point and cannot establish whether such shifts persist or alter behaviour, but the mechanism is straightforward to build into program design and warrants prospective evaluation.(45)

### Theory informed explanatory model for the Positive Running intervention

Figure 2 illustrates how *Positive Running* addresses the intersecting marginalized identities of adolescent girls living with HIV through a framework informed by Intersectionality Theory and grounded in Self Determination Theory.(27,28) The findings underscore the importance of recognizing the multiple, overlapping identities of participants, including being of young age, female gender, living with HIV, and often socioeconomically disadvantaged or orphaned, all of which serve to compound risks of poor health outcomes. These intersecting factors can intensify barriers to participation, underscoring how gendered and structural inequities shape health experiences. As shown in Figure 2, program components promoting autonomy, competence, and relatedness were described by participants as building confidence and self-management. The model also makes visible where the program’s reach ends: components operate at the point of delivery, while the principal constraints participants described operate in households and communities that the program does not enter. This gap between where support is offered and where constraint originates is, we suggest, the central design problem for physical activity programming with adolescent girls in this setting.

**Figure 2.**
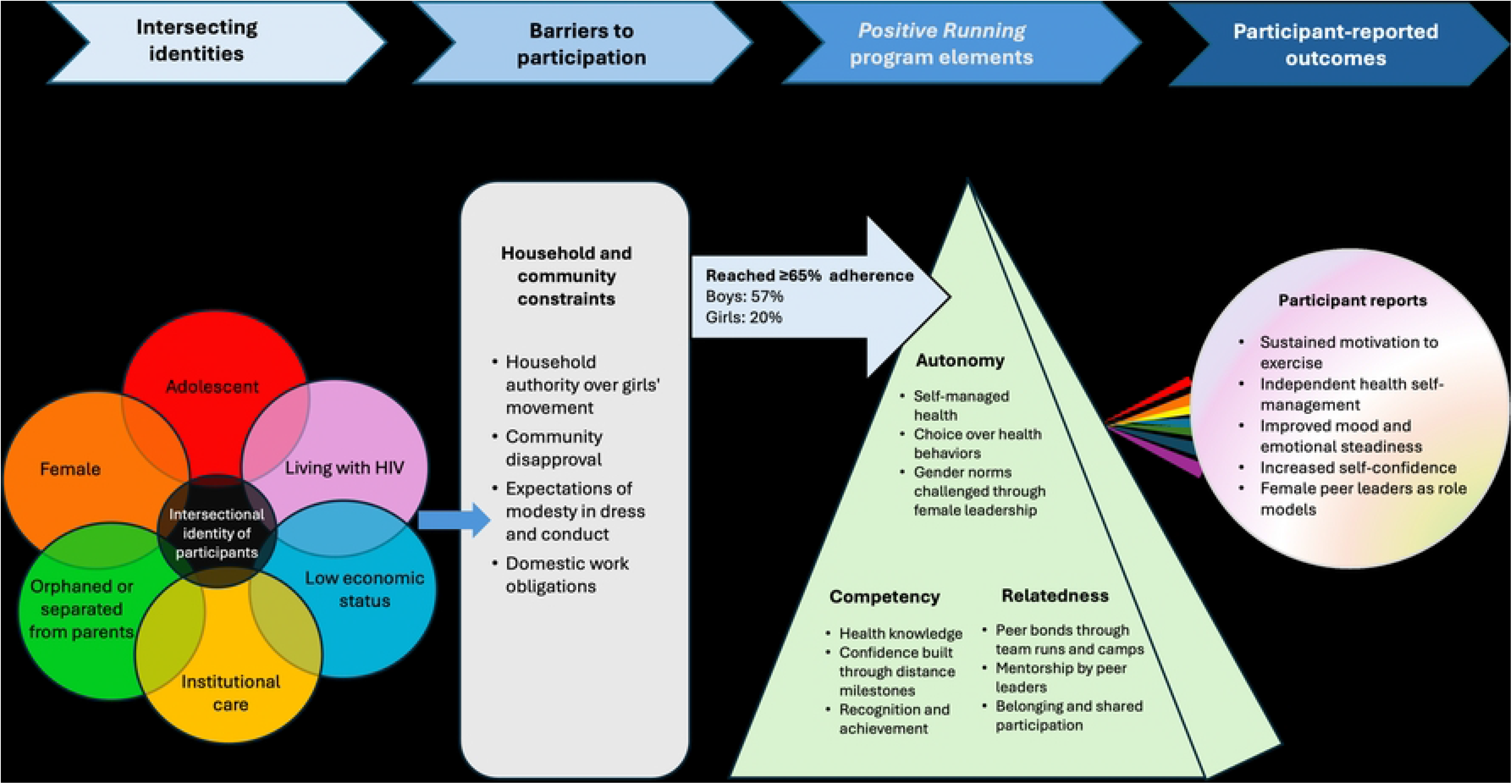
Intersecting identities, barriers, and program elements shaping participation in the Positive Running program

Findings from *Positive Running* reinforce the need for adolescent-centered HIV programs that integrate physical activity, mental health, and gender equity into a cohesive framework. While biomedical adherence and viral suppression remain critical, interventions that nurture autonomy, knowledge-building, and peer connectedness may enhance long-term resilience and retention in care. The peer-led approach is potentially both scalable and sustainable, requiring minimal resources while promoting local ownership and continuity. Scaling this model across community and institutional settings could strengthen India’s adolescent HIV response, particularly for girls and women, who in this evaluation were substantially less able to access the program than the boys and men."

### Convergence and divergence

The adherence gap among boys and girls was closely matched by participants’ accounts of household authority, community disapproval, and competing domestic work, illustrating how the two strands converged on access. However, they diverged on benefit. Participants described the program in largely positive terms, while the quantitative data could not establish whether participation improved mental health or whether less distressed adolescents were simply better able to attend. This divergence is partly structural. Only adolescents who attended could describe what attending offered, which means that young women of greatest interest to this paper are those who could not take part, and are also the least represented in the qualitative data.

### Strengths and limitations

#### Strengths

The convergent mixed-methods design allowed patterns of participation observed quantitatively to be interpreted through participants’ own accounts. The community-based participatory approach, with trained youth investigators with lived experience of HIV contributing to instrument development, data collection, and interpretation, strengthened the cultural and contextual validity of the findings. Gender-disaggregated analysis of physical activity participation in this population is, to our knowledge, rare, and provides important nuance often missing from adolescent physical activity and HIV research.

### Limitations

Several limitations warrant consideration. The cross-sectional design precludes causal inference between program adherence and mental health symptoms. The study was not powered for gender-stratified estimation. With 10 girls meeting the adherence threshold, the female-gender stratified estimate is uninformative, and no sparse-data correction was applied; this estimate should not be interpreted as evidence of differential effect. Adherence was dichotomized at a threshold close to the sample mean, and although sensitivity analyses at alternative thresholds were directionally consistent, dichotomization at any cut-point discards information. Attendance was recorded by peer implementers who also encouraged participation, and was not independently verified. Self-determination theory constructs were interpreted from qualitative accounts rather than measured, and intersectionality was applied as an interpretive frame in a context where participants named gender and age more explicitly than HIV status. Although facilitators were drawn from the program and were familiar to the participants, this familiarity may have encouraged candor about gendered constraints while also making participants less willing to voice criticism of the program itself. Finally, the single geographic focus, the predominance of participants in institutional care, and the absence of a comparison group limit generalizability, although the barriers identified are consistent with findings across diverse cultural contexts.(46)

## CONCLUSION

This evaluation found that *Positive Running*, a peer-led physical activity program for adolescents living with HIV reached girls and young women far less consistently than boys and young men, and that the reasons participants gave lay in household authority, community expectations, and gendered norms operating well before the program’s point of delivery. The disparity was not in benefit received but in the opportunity to participate. This distinction locates the problem where intervention must be directed. Two implications follow. Rather than working with girls and women alone, programs must engage the gatekeepers of girls’ participation, including parents, male household heads, and community leaders. Young men should be part of the solution rather than an obstacle to it: our findings suggest that mixed-gender training can shift attitudes as well as fitness. While the specific estimates reported here reflect a single program in southern India, the analytic insight can be informative in other settings. Where programs are voluntary and delivered in public space, gendered constraints on mobility and time may determine access long before they shape response, and evaluations that measure only outcomes among those who attend will not detect this. Given its grounding in peer leadership and lived experience, the *Positive Running* model offers a culturally resonant framework. Realizing the full potential of this model requires treating the cultural dynamics of gender inequality not as contextual background to intervention design but as a determinant of who the intervention reaches, and therefore a core target of design in itself.

## LIST OF ABBREVIATIONS

APHIV: Adolescents and young adults with perinatally acquired HIV
ART: Antiretroviral therapy
aOR: Adjusted odds ratio
CBPR: Community-based participatory research
CD4: Cluster of differentiation 4
CI: Confidence interval
FGD: Focus group discussion
GAD: Generalized anxiety disorder
GAD-7: 7-item Generalized Anxiety Disorder scale
HIV: Human immunodeficiency virus
IDI: In-depth interview
IQR: Interquartile range
OR: Odds ratio
PHQ-9: 9-item Patient Health Questionnaire
PRP: Positive Running Program
SD: Standard deviation
SDG: Sustainable Development Goal
SDT: Self-Determination Theory
VL: Viral load
WHO: World Health Organization
YRGCARE: Y.R. Gaitonde Centre for AIDS Research and Education

## Data Availability

The de-identified quantitative dataset supporting the findings reported in this article is available as supplementary material. Sensitive information such as dates, study site and institution identifiers have been removed in order to protect the confidentiality of the participant population. For the qualitative data, the study codebook and illustrative excerpts supporting the reported themes are available as supplementary materials. Full interview and focus group transcripts cannot be publicly released because they cannot be adequately de-identified and public sharing falls outside the scope of the consent and assent provided by participants. Requests for access to additional de-identified qualitative excerpts may be directed to the Institutional Ethics Committee of the Y.R. Gaitonde Centre for AIDS Research and Education (YRGCARE), Chennai, India, and/or to the corresponding author. Access may be granted in accordance with the approved study protocol and applicable Indian data protection requirements.

